# A predictive model for left ventricular hypertrophy in hypertensive children

**DOI:** 10.1101/2025.10.15.25338126

**Authors:** Yang Liu, Yaxi Cui, Yao Lin, Tong Zheng, Min Bao, Yanyan Liu, Lin Shi

## Abstract

**Background:** As current electrocardiographic (ECG) criteria are of low diagnostic value compared with echocardiography (ECHO) for LVH, establishing a more effective ECG predictive model is necessary.

**Objective:** To develop and validate model to improve the diagnostic capability of ECG for LVH in pediatric primary hypertension.

**Methods:** A retrospective study of 502 hypertensive children were recruited in the study between January 2019 and December 2024. The cohort were randomly divided into train (n=402) and test sets (n=100) with a proportion of 8:2. LVH was diagnosed using ECHO criteria. A total of 22 ECG parameters were evaluated. A predictive nomogram was developed using least absolute shrinkage and selection operator (LASSO) and multivariate logistic regression.

**Results:** LVH was identified in 117 (29.1%) of the training set and 29 (29.0%) of the test set. Body mass index (BMI), R_I_ + S_V4_, and S_D_ + S_V4_ were identified as independent predictors of LVH. The nomogram model showed good performance, with an area under the curve (AUC) of 0.822 in the training set and 0.803 in the test set. Calibration curves and Hosmer-Lemeshow test indicated good agreement between predicted and actual probabilities. DCA demonstrated clinical usefulness. The model outperformed previous models, as confirmed by NRI and IDI.

**Conclusions:** The nomogram model incorporating BMI, R_I_ + S_V4_, and S_D_ + S_V4_ significantly improves the ECG diagnosis of LVH in pediatric primary hypertension. The model offers a reliable tool for the early detection of LVH in hypertensive children.

## Introduction

Hypertension is a key risk factor for cardiovascular disease in adults.^1^ Elevated blood pressure throughout childhood or adolescence raises the likelihood of cardiovascular events in adulthood.^2^ Contrary to adult hypertension, pediatric hypertension rarely results in cardiovascular events; however, subclinical target organ damage (TOD) is frequently observed.^3,4^ That’s why TOD is frequently utilized as the endpoint event for assessing cardiovascular risk in children with primary hypertension. Left ventricular hypertrophy (LVH) is the most significant indication of hypertensive TOD which is currently used to indicate the risk of cardiovascular complications in hypertensive children in adulthood.^5,6^ Therefore, early identification of LVH is of great significance for the early prevention and treatment of hypertensive cardiovascular diseases in children. Echocardiography (ECHO) has been shown to be an accurate predictor of LVH;^7^ However, electrocardiography (ECG) is less expensive, quicker, and more easily performed. The 12-lead ECG is regarded as the initial diagnostic indicator for LVH, there are many ECG indicators that can be used to identify LVH in hypertensive patients, yet these ECG indicators are often of low diagnostic value compared with ultrasonic indicators, and are often ignored by clinicians due to their complex diversity.^8^ Several ECG criteria, such as the Sokolow-Lyon index, Cornell voltage or Cornell voltage duration product, and RavL, are available to assess LVH in adult hypertensive patients.^9^ However the diagnostic performance of ECG criteria in children with primary hypertension remains under investigation.^10^ Therefore, it is necessary to compare the clinical application value of various common ECG indicators, and then optimize the comprehensive application of ECG indicators. The objective of the present study was designed to evaluate the diagnostic value of various ECG criteria for LVH in children with primary hypertension, using ECHO as the gold standard in the diagnosis of LVH. Based on these findings, we aimed to develop an easy-to-perform, highly sensitive, and specific predictive model to improve the diagnostic ability of ECG criteria for LVH.

## Methods

### Study Design and subjects

This was a retrospective study in a single center. The study included 502 pediatric patients, aged 6-17 years, including 400 males (79.68%) and 102 females (20.32%), who were first diagnosed with primary hypertension hospitalized in the Capital Center for Children’s Health, Capital Medical University (Beijing, China), between January 2019 and December 2024. All enrolled patients were randomly divided into train (n = 402) and test sets (n = 100) with a proportion of 8:2.

### Ethics

This study had approval from the Ethic Committee of the Capital Center for Children’s Health, Capital Medical University (No: SHERLL2025011). The parents/guardians of all patients provided signed informed consent prior to enrolment.

### Patient selection criteria

The diagnosis was made following the indications proposed in the Chinese guidelines for the diagnosis and management of hypertension in children and adolescents.^11^ All blood pressure measurements were performed using the auscultation method as recommended.^12^ Hypertension diagnosis was reflected by average systolic blood pressure (SBP) and/or diastolic blood pressure (DBP) beyond the 95th percentiles of the auscultation measurement, after adjustments for gender, age, and height. Stages 1 and 2 hypertension were diagnosed with BP<99th and≥99th percentiles, respectively, plus 5 mmHg.

Exclusion criteria were secondary hypertension associated with kidney, vascular, endocrine, or central nervous system disease; Patients with other systemic diseases such as respiratory diseases, acute or chronic liver diseases, hyperthyroidism, diabetes, and immune system diseases; Primary hypertension previously treated with antihypertensives.

### Echocardiography

LVH was assessed by echocardiography. A Philips iE33 Ultrasound System (Philips Healthcare, USA) was utilized to measure left ventricular internal dimension (LVIDd), interventricular septal thickness (IVST), and left ventricular posterior wall thickness (LVPWT) at the end of diastole. LVM was derived as 1.04×0.8× ((LVIDd + IVST + LVPWT)^3^ – LVIDd^3^) +0.6; LVM index (LVMI) was determined as LVM/height^2.7^. Relative left ventricular wall thickness (RWT) was derived as (IVST+LVPWT)/LVIDd.^7^ For the diagnosis of LVH, (a) LVMI≥37.08 g/m^2.7^ in males, LVMI≥34.02 g/m^2.7^ in females, respectively; (b) RWT>0.36 in either males or females. Fulfillment of either criterion (a) or (b) was defined as LVH.^13,14^ Experienced cardiologists performed echocardiographic assessments to ensure unbiased results.

### Electrocardiography (Different electrocardiographic criteria for LVH)

An ECG was performed for every patient during the same hospitalization period when the ECHO was obtained. A standard 12-lead ECG was conducted by trained technicians at rest using the DMS 300-BTT02 ECG workstation (DMS, Beijing, China), with a velocity and voltage regulation of 25 mm/s and 1 mV/10 mm, respectively. The voltages of the R, S, and Q waves, as well as the QRS duration, were measured in all leads. A total of 22 ECG parameters for diagnosing LVH in children were evaluated^15–20^, which were calculated from the automatically measured ECG waveforms and mainly used in clinical practice(Table 1), with an additional person responsible for the verification and correction to ensure the accuracy and stability of all electrocardiogram data.

**Table 1.**
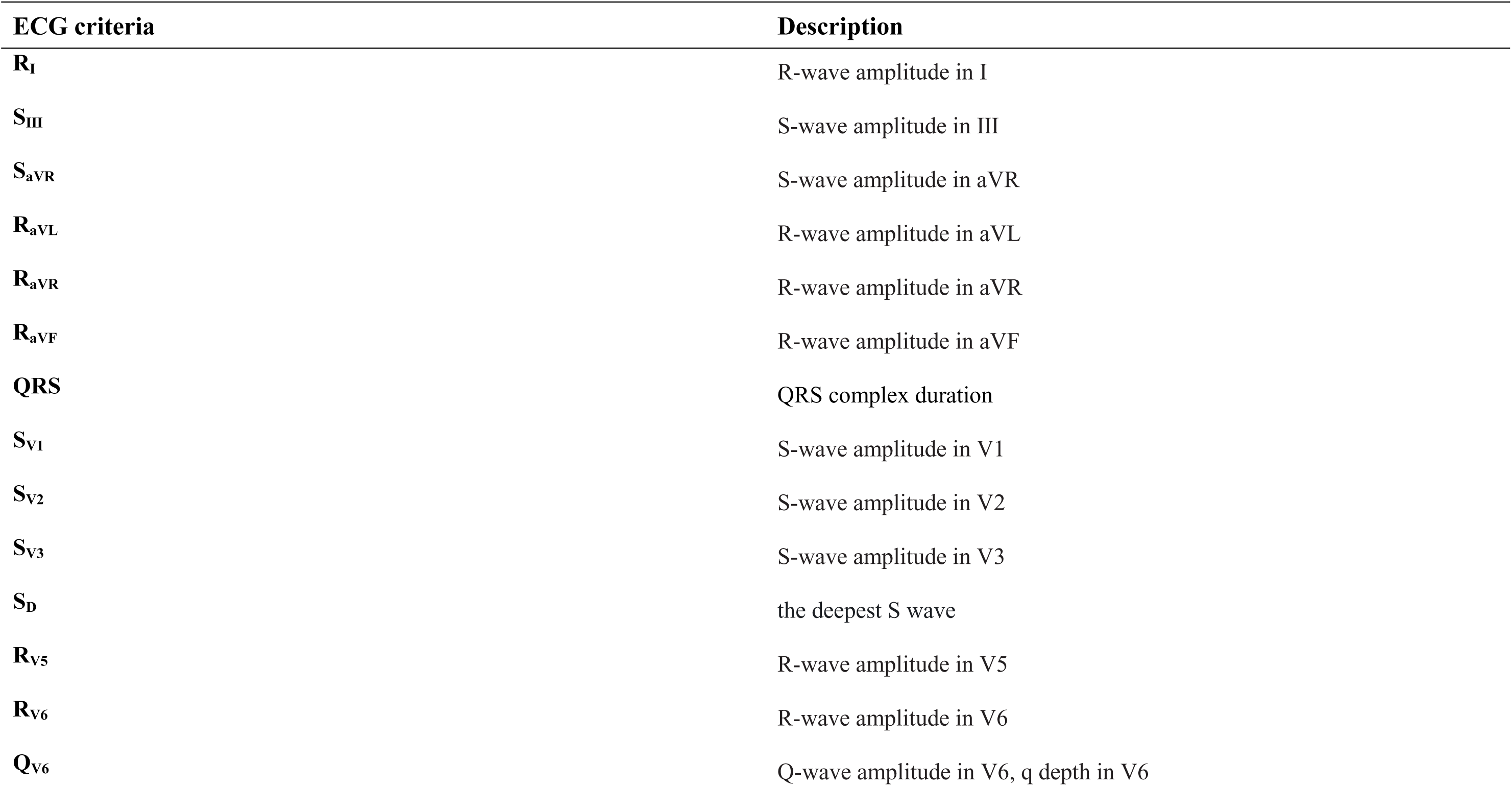
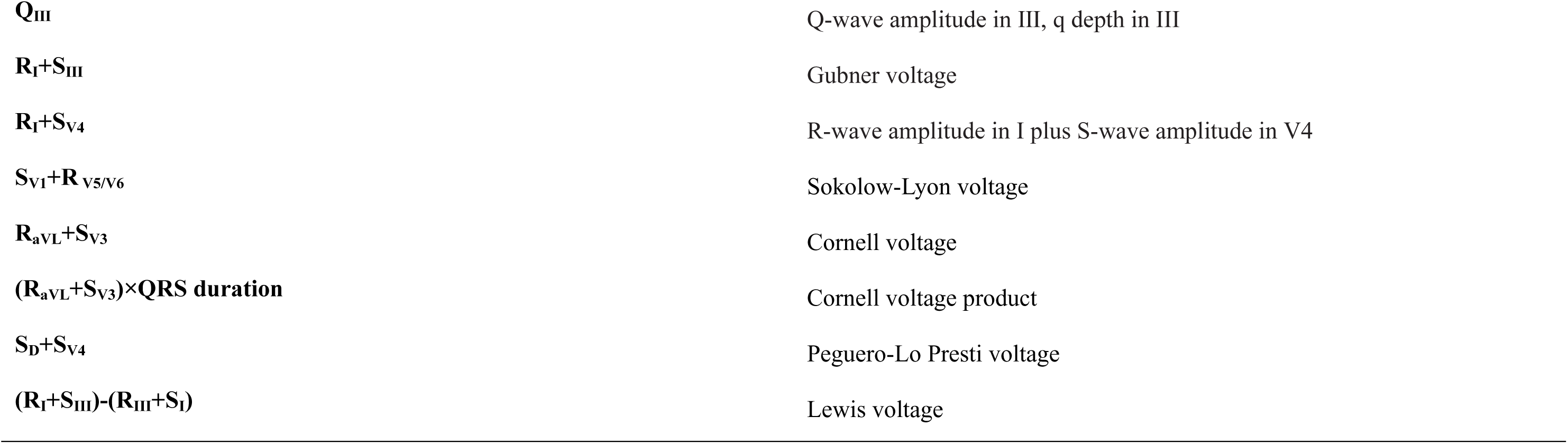
Evaluated ECG parameters for LVH in children.

| **ECG criteria** | **Description** |
| --- | --- |
| **R<sub>I</sub>** | R-wave amplitude in I |
| **S<sub>III</sub>** | S-wave amplitude in III |
| **S<sub>aVR</sub>** | S-wave amplitude in aVR |
| **R<sub>aVL</sub>** | R-wave amplitude in aVL |
| **R<sub>aVR</sub>** | R-wave amplitude in aVR |
| **R<sub>aVF</sub>** | R-wave amplitude in aVF |
| **QRS** | QRS complex duration |
| **S<sub>V1</sub>** | S-wave amplitude in V1 |
| **S<sub>V2</sub>** | S-wave amplitude in V2 |
| **S<sub>V3</sub>** | S-wave amplitude in V3 |
| **S<sub>D</sub>** | the deepest S wave |
| **R<sub>V5</sub>** | R-wave amplitude in V5 |
| **R<sub>V6</sub>** | R-wave amplitude in V6 |
| **Q<sub>V6</sub>** | Q-wave amplitude in V6, q depth in V6 |
| $Q_{III}$ | Q-wave amplitude in III, q depth in III |
| $R_I+S_{III}$ | Gubner voltage |
| $R_I+S_{V4}$ | R-wave amplitude in I plus S-wave amplitude in V4 |
| $S_{V1}+R_{V5/V6}$ | Sokolow-Lyon voltage |
| $R_{aVL}+S_{V3}$ | Cornell voltage |
| $(R_{aVL}+S_{V3}) \times \text{QRS duration}$ | Cornell voltage product |
| $S_D+S_{V4}$ | Peguero-Lo Presti voltage |
| $(R_I+S_{III})-(R_{III}+S_I)$ | Lewis voltage |

### Statistical Analysis

R software (version: 4.4.3, http://www.R-project.org). Two-sided P<0.05 indicated statistical significance.

Within a normal distribution, measurement data were expressed as mean ± standard deviation, while those with skewed distribution were represented by median and interquartile range *M* (*Q_1_*, *Q_3_*). Categorical variables (percentage (%)) were compared by the *χ*^2^-test. The independent samples t-test or Mann–Whitney U-test was used for intergroup comparisons of continuous variables and the Chi-square test was used for categorical variables.

Variables with significant differences (P < 0.10) between the LVH and normal geometry group in pediatric patients with primary hypertension were preliminarily screened by least absolute shrinkage and selection operator (LASSO) regression in the train set. LASSO analysis shrunk the regression coefficient of variables to zero by a penalized coefficient of lambda. Variables without zero regression coefficients were selected to enter into a multivariable logistic regression using a backward stepwise regression approach. Predictors with statistical significance were retained as the final independent variables for model construction.

A nomogram was developed based on the final logistic regression model to illustrate the model results. Discrimination of models was assessed using the receiver operating characteristic (ROC) curve. The area under the curve (AUC) for 0.7 or more indicated good discrimination. The prediction accuracy was assessed by calibration plots and the Hosmer–Lemeshow. The clinical utility was estimated by decision curve analysis (DCA). The nomogram was validated by the test set.

To compare the predictive performance of the developed model with that of previously published models, we calculated and compared the AUC, net reclassification improvement (NRI), and integrated discrimination improvement (IDI) indices.

## Results

### Patients’ characteristics

A total of 502 hypertensive children, aged 13 (12, 14) years old, were recruited in the study, including 400 males (79.68%) and 102 females (20.32%) between January 2019 and December 2024. All enrolled patients were randomly divided into train (n = 402) and test sets (n = 100) with a proportion of 8:2 (Figure 1). There was no significant difference in clinical variables and ECG parameters between train and test sets (Table 2). LVH was observed in 117 (29.1%) patients in the train test and 29 (29.0%) in the test set.

**Figure 1.**
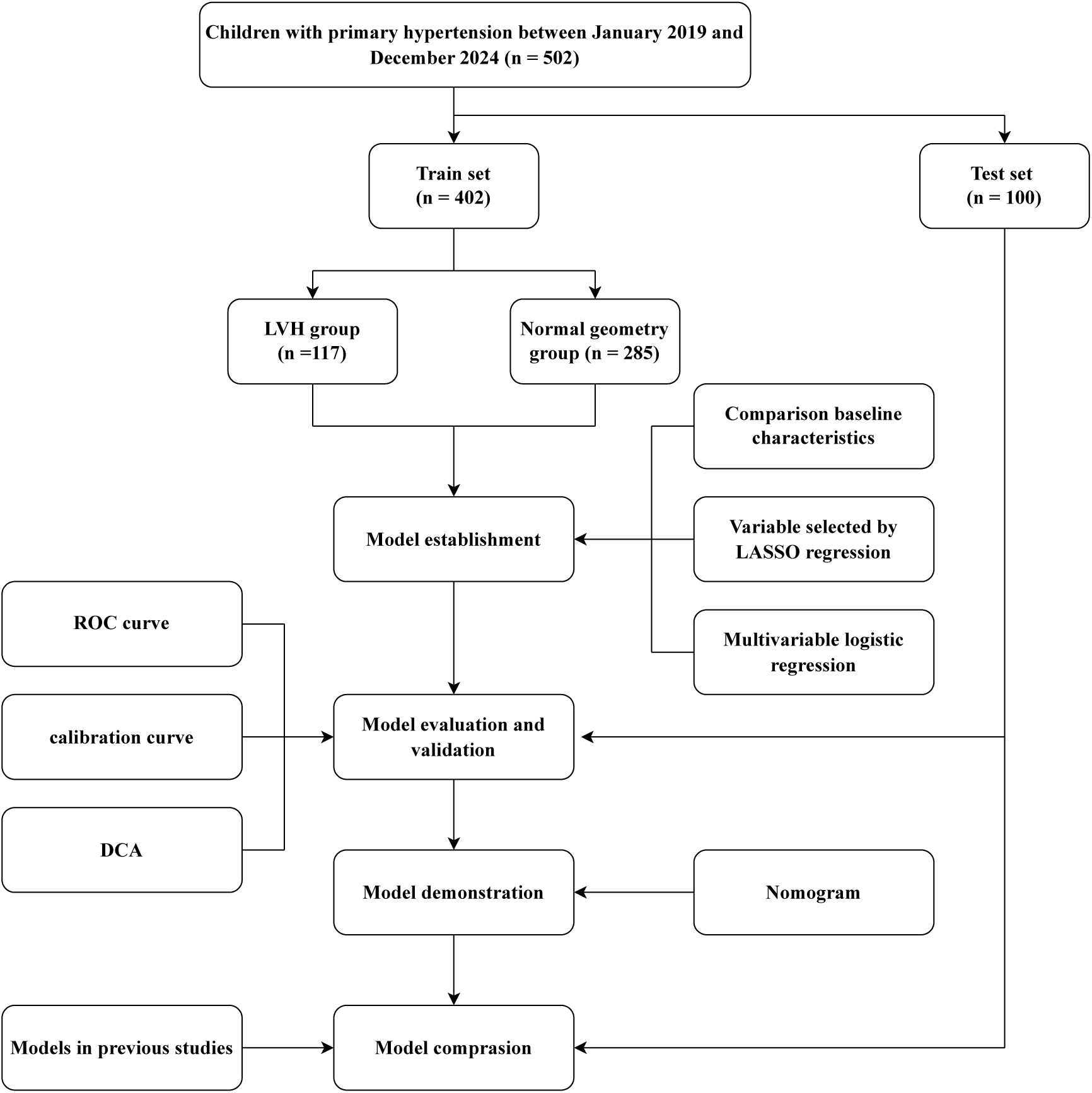
The flowchart of the research. LVH, left ventricular hypertrophy; LASSO, least absolute shrinkage and selection operator; ROC, receiver operating characteristic; DCA, decision curve analysis.

**Table 2.** Comparison of characteristics between train and test sets.

| Variables | All (n=502) | Train (n=402) | Test (n=100) | P value |
| --- | --- | --- | --- | --- |
| **LVH (%)** | 146 (29.08%) | 117 (29.10%) | 29 (29.00%) | 1.000 |
| **Demographic parameters** |  |  |  |  |
| **Age (years)** | 13.00 (12.00,14.00) | 13.00 (12.00,14.00) | 13.00 (12.00,14.00) | 0.872 |
| **Sex (%)** |  |  |  | 0.181 |
| **Male** | 400 (79.68%) | 315 (78.36%) | 85 (85.00%) |  |
| **Female** | 102 (20.32%) | 87 (21.64%) | 15 (15.00%) |  |
| **Obesity (%)** | 378 (75.30%) | 305 (75.87%) | 73 (73.00%) | 0.641 |
| **Height (cm)** | 1.71 (1.63,1.78) | 1.70 (1.63,1.78) | 1.72 (1.64,1.76) | 0.718 |
| **Weight (kg)** | 80.00 (68.00,92.97) | 80.00 (68.00,92.90) | 80.00 (68.00,93.80) | 0.657 |
| **BMI (kg/m<sup>2</sup>)** | 27.43 (24.30,30.70) | 27.30 (24.30,30.58) | 27.76 (24.35,31.45) | 0.494 |
| **Stage (%)** |  |  |  | 0.567 |
| **1** | 194 (38.65%) | 152 (37.81%) | 42 (42.00%) |  |
| **2** | 308 (61.35%) | 250 (62.19%) | 58 (58.00%) |  |
| **Duration (%)** |  |  |  | 0.272 |
| **<6 month** | 265 (52.79%) | 209 (51.99%) | 56 (56.00%) |  |
| **6-12 month** | 82 (16.33%) | 71 (17.66%) | 11 (11.00%) |  |
| **>12 month** | 155 (30.88%) | 122 (30.35%) | 33 (33.00%) |  |
| **History (%)** | 391 (77.89%) | 312 (77.61%) | 79 (79.00%) | 0.869 |
| **SBP (mmHg)** | 131.00 (124.00,137.00) | 130.50 (123.25,136.00) | 132.00 (125.00,138.25) | 0.164 |
| **DBP (mmHg)** | 72.00 (68.00,77.00) | 72.00 (68.00,77.00) | 72.00 (68.00,76.00) | 0.615 |
| **ECG parameters** |  |  |  |  |
| **R<sub>I</sub>** | 0.65 (0.49,0.85) | 0.64 (0.49,0.85) | 0.66 (0.49,0.88) | 0.587 |
| **R<sub>avR</sub>** | 0.06 (0.02,0.18) | 0.06 (0.02,0.18) | 0.08 (0.02,0.22) | 0.152 |
| **R<sub>avF</sub>** | 0.94 (0.64,1.25) | 0.94 (0.65,1.25) | 0.94 (0.61,1.25) | 0.809 |
| **R<sub>v5</sub>** | 1.51 (1.21,1.82) | 1.50 (1.23,1.83) | 1.52 (1.15,1.81) | 0.780 |
| **R<sub>v6</sub>** | 1.27 (1.03,1.52) | 1.27 (1.04,1.51) | 1.27 (0.93,1.53) | 0.603 |
| **QRS** | 96.00 (90.00,104.00) | 96.00 (90.00,104.00) | 96.00 (89.50,104.00) | 0.781 |
| **S<sub>III</sub>** | 0.00 (0.00,0.14) | 0.00 (0.00,0.13) | 0.02 (0.00,0.16) | 0.169 |
| **S<sub>aVR</sub>** | 0.58 (0.00,1.00) | 0.62 (0.00,1.01) | 0.00 (0.00,0.94) | 0.406 |
| **S<sub>v1</sub>** | 0.91 (0.61,1.30) | 0.92 (0.62,1.30) | 0.89 (0.60,1.25) | 0.608 |
| **S<sub>v2</sub>** | 1.45 (1.11,1.86) | 1.44 (1.09,1.86) | 1.49 (1.15,1.86) | 0.569 |
| **S<sub>v3</sub>** | 0.76 (0.46,1.07) | 0.76 (0.45,1.07) | 0.76 (0.54,1.03) | 0.729 |
| **S<sub>D</sub>** | 1.52 (1.21,1.90) | 1.50 (1.21,1.90) | 1.58 (1.24,1.88) | 0.622 |
| **Q<sub>III</sub>** | 0.04 (0.00,0.14) | 0.05 (0.00,0.14) | 0.03 (0.00,0.14) | 0.263 |
| **Q<sub>v6</sub>** | 0.02 (0.00,0.06) | 0.02 (0.00,0.06) | 0.01 (0.00,0.04) | 0.103 |
| **S<sub>v1</sub>+R<sub>v5</sub>** | 2.46 (1.96,2.94) | 2.46 (1.98,2.94) | 2.46 (1.92,2.95) | 0.680 |
| **S<sub>v1</sub>+R<sub>v6</sub>** | 2.18 (1.76,2.68) | 2.18 (1.77,2.69) | 2.17 (1.75,2.66) | 0.561 |
| **R<sub>avL</sub>+S<sub>v3</sub>** | 1.03 (0.70,1.38) | 1.02 (0.68,1.38) | 1.06 (0.78,1.38) | 0.405 |
| **(R<sub>I</sub>+S<sub>III</sub>)-(R<sub>III</sub>+S<sub>I</sub>)** | -0.18 (0.83) | -0.19 (0.82) | -0.12 (0.88) | 0.456 |
| **R<sub>avL</sub>** | 0.22 (0.12,0.36) | 0.22 (0.12,0.35) | 0.23 (0.13,0.38) | 0.408 |
| **R<sub>I</sub>+S<sub>III</sub>** | 0.73 (0.52,0.97) | 0.72 (0.52,0.95) | 0.76 (0.51,1.02) | 0.383 |
| **R<sub>I</sub>+S<sub>v4</sub>** | 1.11 (0.86,1.40) | 1.10 (0.84,1.41) | 1.13 (0.90,1.37) | 0.494 |
| **(R<sub>aVL</sub>+S<sub>v3</sub>)×QRS duration** | 97.29 (68.00,134.64) | 95.00 (66.27,134.19) | 101.65 (70.55,137.20) | 0.466 |
| **S<sub>D</sub>+S<sub>v4</sub>** | 1.95 (1.54,2.45) | 1.94 (1.53,2.49) | 1.96 (1.67,2.36) | 0.687 |

### Comparison baseline characteristics between LVH group and normal geometry group in children with primary hypertension in train set

Of all cases in train set, 117 (29.1%) had LVH as defined by the pediatric ECHO criteria, and 285 (70.9%) had normal cardiac geometry. Demographic characteristics, and ECG parameters between the two groups are shown in Table 3. Compared to normal cardiac geometry group, the LVH group had a significantly higher SBP, weight, BMI, R_I_, R_aVR_, QRS, S_V1_, S_V2_, S_V3_, S_D_, Q_Ⅲ_, S_V1_ plus R_V6_, R_aVL_ plus S_V3_, R_aVL_, R_Ⅰ_ plus S_Ⅲ_, R_Ⅰ_ plus S_V4_, (R_aVL_ plus S_V3_) multiplied by QRS complex duration, S_D_ plus S_V4_. (Table 3).

**Table 3.**
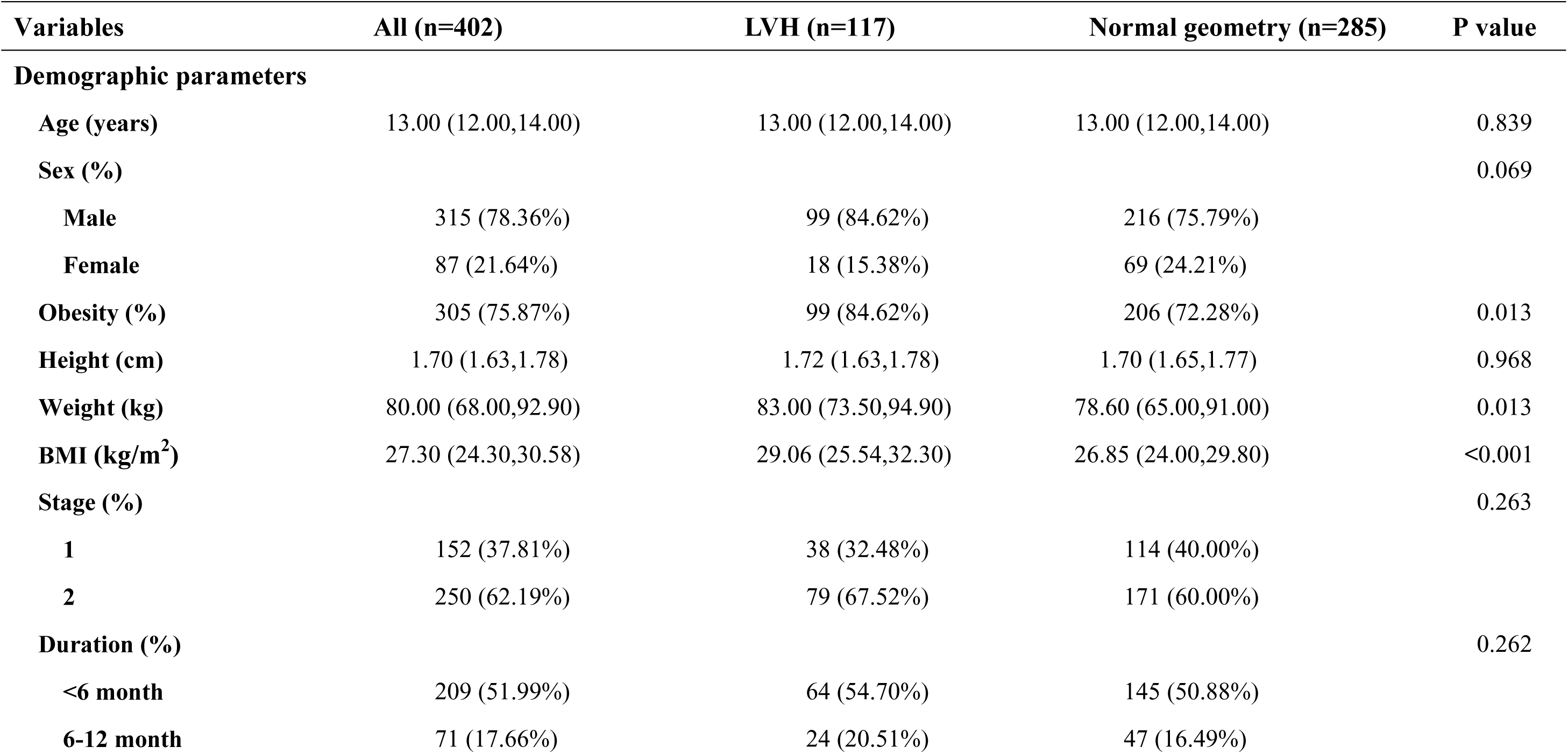
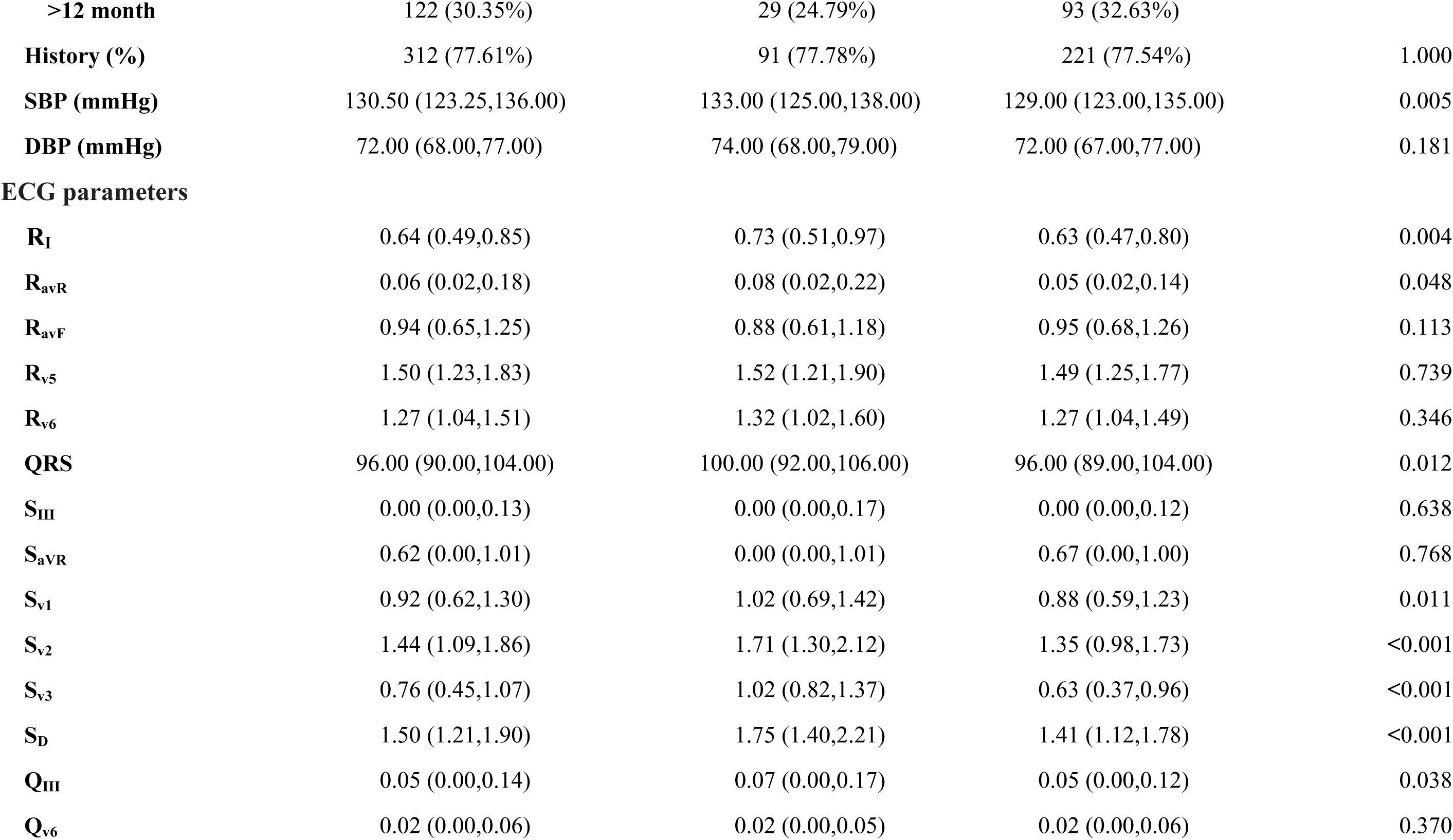
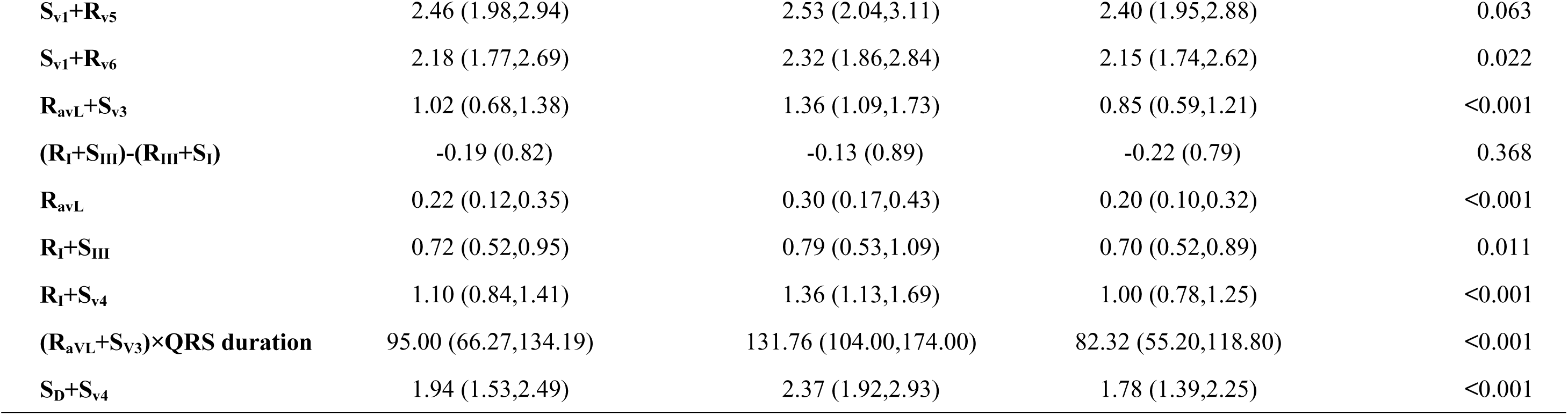
Comparison baseline characteristics between LVH group and normal geometry group in children with primary hypertension in train set.

### Predictors of LVH

A total of 21 variables with a *P* value < 0.1 in the univariate comparison (Table 3) between the LVH group and the normal cardiac geometry group were entered into the LASSO regression. Using 10-fold cross-validation, the optimal penalty parameter was determined based on the 1-standard error criterion (λ.1se = 0.0412), which resulted in four predictors with nonzero coefficients: BMI, R_aVL_ + S_V3_, R_I_ + S_V4_, and S_D_ + S_V4_ (Figure 2). These four variables were subsequently entered into a multivariable binary logistic regression. The final model retained three independent predictors of LVH: BMI (OR:1.117; 95%CI: 1.052–1.185), R_I_ + S_V4_ (OR: 5.317; 95%CI: 2.379–11.883), and S_D_ + S_V4_ (OR: 4.417; 95%CI: 2.700–7.226) (Table 4). The logistic regression equation was as follows: Logit (*P*) = –9.199 + 0.110 × BMI + 1.671× (R_Ⅰ_+ S_V4_) + 1.485 × (S_D_ + S_V4_).

**Figure 2.**
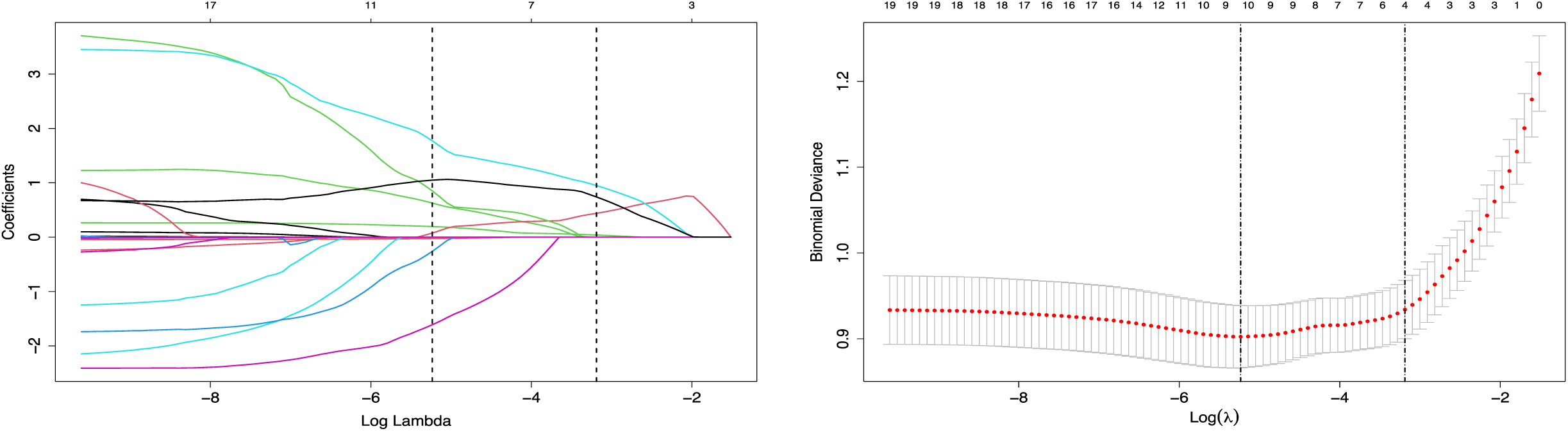
LASSO regression for identifying predictors of LVH in children with primary hypertension. (A) In LASSO regression, the penalty term λ is introduced; as λ increases, stronger shrinkage is applied, driving the coefficients of many features toward zero. By selecting features with nonzero coefficients, the model retains those variables with the greatest predictive ability for the outcome, thereby simplifying the model and improving its performance. (B) Cross-validation curve for the penalty parameter λ. The x-axis represents log(λ), and the y-axis indicates the mean squared error (MSE). The left dashed line corresponds to the λ value that yields the minimum MSE, while the right dashed line represents the largest λ within one standard error of the minimum MSE. LVH, left ventricular hypertrophy; LASSO, least absolute shrinkage and selection operator.

**Table 4.** Multivariate logistic regression analysis of predictors selected by LASSO regression in the development set.

| **Variables** | **Coefficient** | **SE** | **OR** | **95% CI for OR** | **P** |
| --- | --- | --- | --- | --- | --- |
| **BMI** | 0.11 | 0.03038 | 1.117 | 1.117 (1.052–1.185) | <0.001 |
| **$R_I+S_{v4}$** | 1.671 | 0.41033 | 5.317 | 5.317 (2.379–11.883) | <0.001 |
| **$S_D+S_{v4}$** | 1.485 | 0.25112 | 4.417 | 4.417 (2.700–7.226) | <0.001 |

### Establishment of the nomogram model

The nomogram was developed based on three independent predictors identified in the multivariable logistic regression model: body mass index (BMI), R_I_ + S_V4_, and S_D_ + S_V4_. (Figure 3). Each predictor is assigned a score according to its contribution to the model, and the sum of these scores corresponds to the predicted probability of LVH. For example, in patient ID6, the BMI, R_I_ + S_V4_, and S_D_ + S_V4_ values yielded a total score of 79.6 points, which translated to an estimated 12.7% probability of LVH.

**Figure 3.**
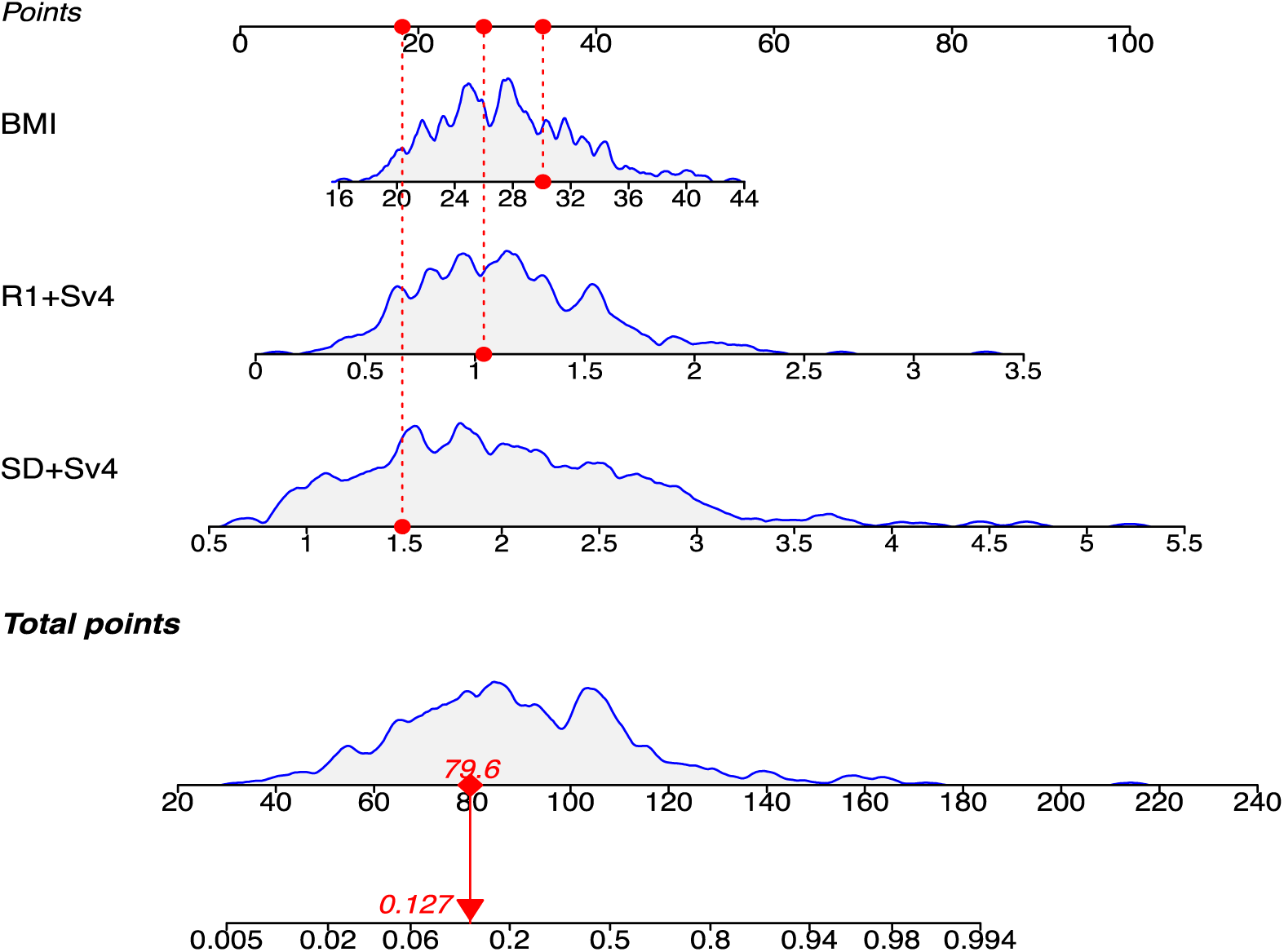
Nomogram for predicting LVH in children with primary hypertension based on electrocardiographic data. The nomogram was developed using three independent predictors identified in the multivariable logistic regression model: BMI, R_I_ + S_V4_, and S_D_ + S_V4_. As an illustrative example, for a child with a BMI of 22.2kg/m^2^, R_I_ + S_V4_ of 0.4, and S_D_ + S_V4_ of 0.66, the individual scores for each predictor sum to a total of 79.6 points, which corresponds to an estimated probability of 12.7% for developing LVH. BMI, body mass index; LVH, left ventricular hypertrophy.

### Evaluation and validation of the nomogram model

The predictive model demonstrated good discriminative ability in both the train and test sets. In the train set, the AUC was 0.822 (95% CI: 0.777–0.867), while in the test set, the AUC was 0.803 (95% CI: 0.713–0.893) (Figure 4). Comparison of the AUCs between the two cohorts using DeLong’s test revealed no statistically significant difference (AUC difference = 0.018, P = 0.719), suggesting that the model’s discriminative performance was stable across datasets.

**Figure 4.**
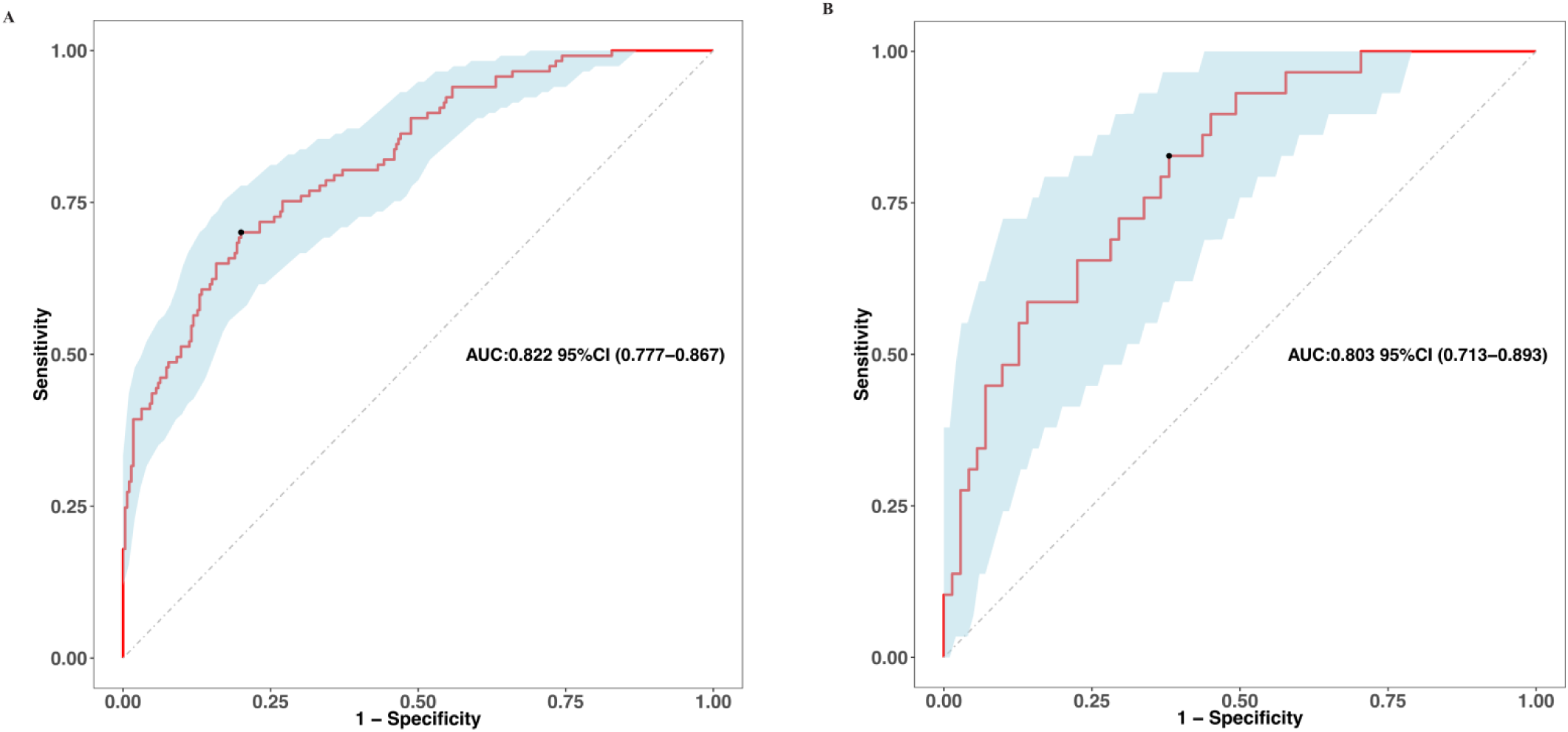
ROC curve analysis of nomogram model of LVH in children with primary hypertension. (A) ROC curve analysis for the train set; (B) ROC curve analysis for the test set. LVH, left ventricular hypertrophy

The calibration curve analysis demonstrated a good alignment between the calibration and standard curves in the train and test sets (Figure 5). The Hosmer-Lemeshow test demonstrated a strong consistency between the predicted and the observed LVH in train set (*χ2* = 7.6879, P = 0.4645) and test set (*χ2* = 2.7444, P = 0.9494). The DCA demonstrated that using the nomogram model can significantly enhance clinical benefits (Figure 6).

**Figure 5.**
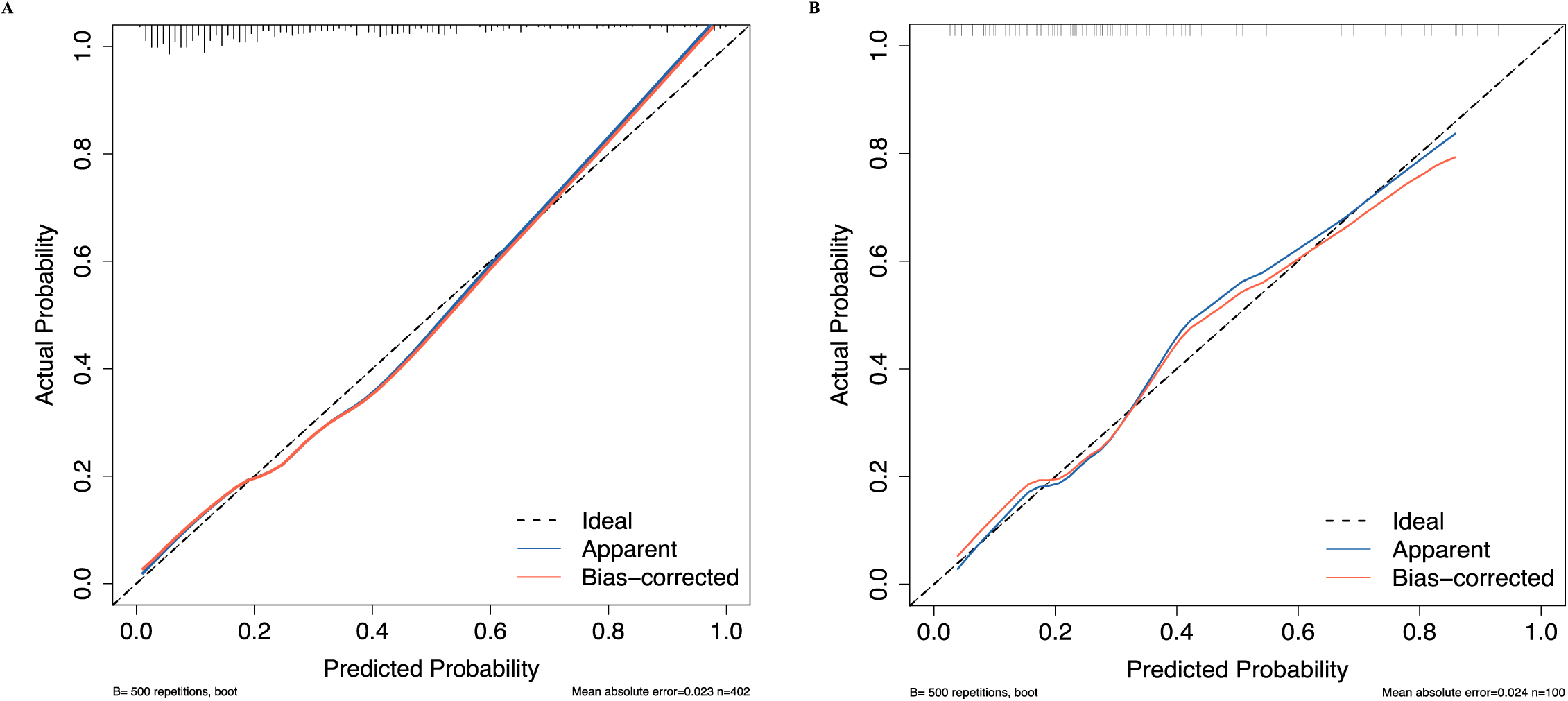
Calibration curve analysis of nomogram model of LVH in children with primary hypertension. (A) Calibration curve analysis in the train set; (B) Calibration curve analysis in the test set. LVH, left ventricular hypertrophy.

**Figure 6.**
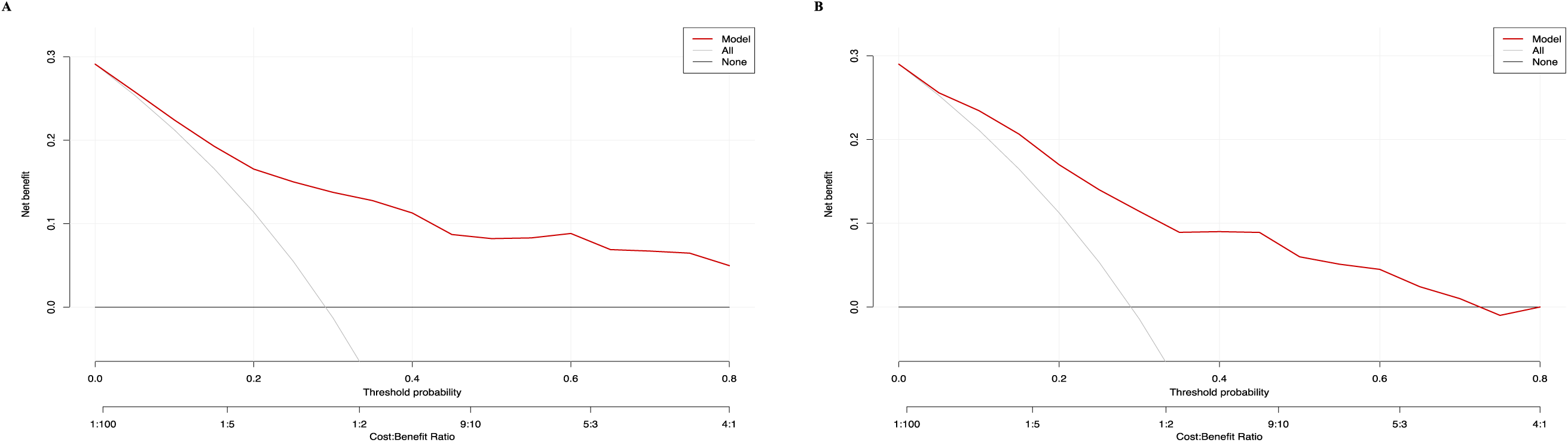
DCA of nomogram model of LVH in children with primary hypertension. (A) DCA for the train set; (B) DCA for the test set. DCA, decision curve analysis. LVH, left ventricular hypertrophy

In the train set, the predictive model achieved an accuracy of 77.1% (95% CI: 72.7%–81.1%) and a Cohen’s kappa coefficient of 0.4746. In the test cohort, the model showed an accuracy of 73.0% (95% CI: 63.2%–81.4%) and a kappa coefficient of 0.3638. Both indicated moderate agreement between predictions and actual outcomes. The sensitivity and specificity were 70.0% and 80.0% in train set, while 58.6% and 78.9% in test set, respectively. McNemar’s test revealed a statistically significant difference between predicted and observed classifications (P = 0.029 in train test and P = 0.700 in test set).

### Model comparison with previous studies

When compared with two published models common used by clinical doctors, our model achieved a higher AUC (0.822, 95% CI: 0.777–0.867) than Model B (0.542, 95% CI: 0.506–0.578) and Model C (0.515, 95% CI: 0.498–0.532) (Figure 7A) in train set. In the test set, Model A similarly outperformed Model B (0.478, 95% CI: 0.418–0.538) and Model C (0.500, 95% CI: 0.500–0.500) (Figure 7B).

**Figure 7.**
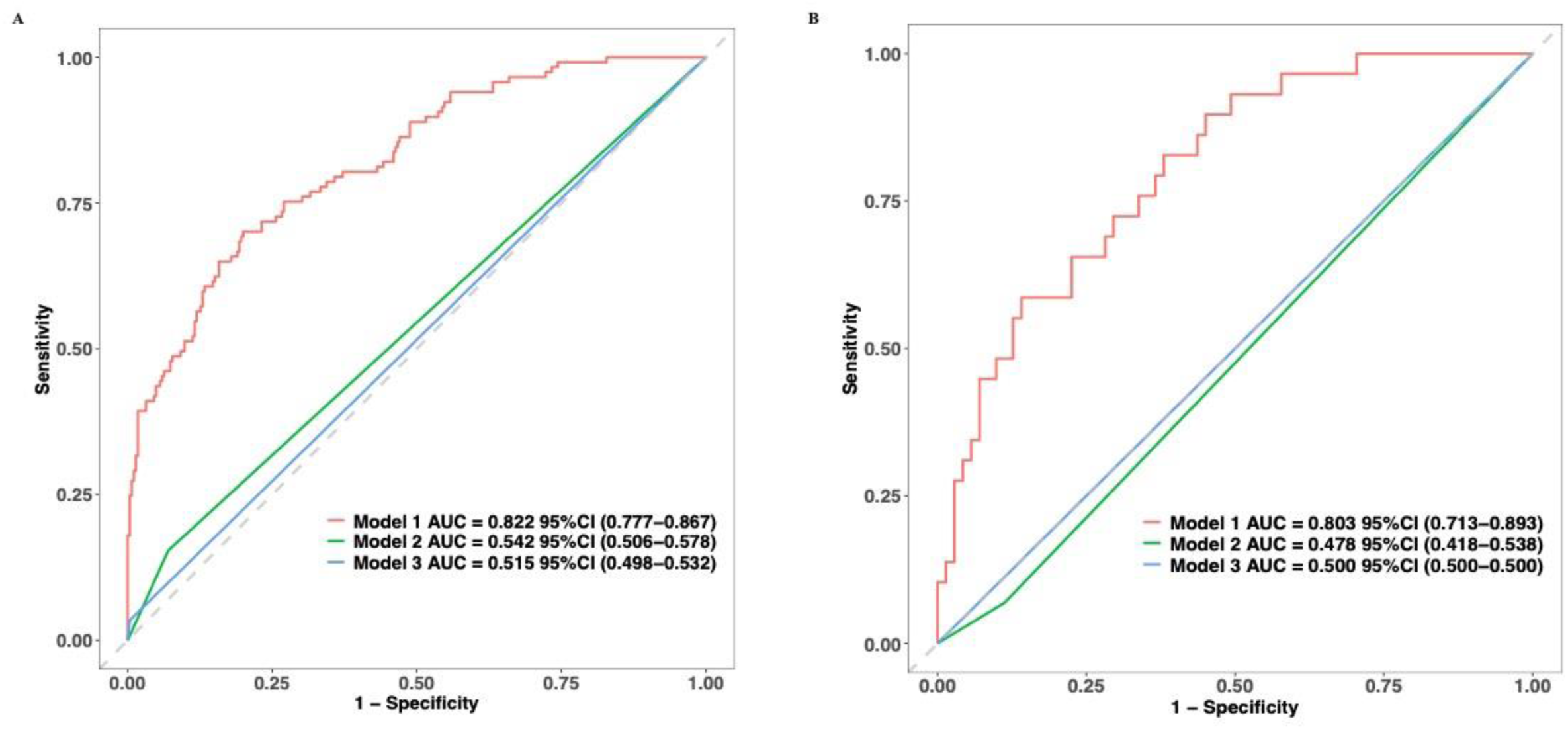
Model comparison between nomogram model with previous models. (A) Model comparison in train set; (B) Model comparison in test set.

NRI and IDI analyses further confirmed the superior performance of the proposed model. In the train set, the categorical NRI was 0.403 (95% CI: 0.307–0.499, P < 0.001), and the IDI was 0.294 (95% CI: 0.238–0.350, P < 0.001). Similar results were observed in test set, with a categorical NRI of 0.372 (95% CI: 0.276–0.468, P < 0.001), and an IDI of 0.295 (95% CI: 0.242–0.348, P < 0.001).

## Discussion

The early phase of hypertensive cardiac injury in children, predominantly manifested as LVH, is a critical intermediate stage in the progression of cardiac damage from childhood to adulthood, thereby holding substantial assessment value.^21^

ECG is the only diagnostic tool that can directly record the electrical activity of the heart. Since electrical activity precedes myocardial contraction, it complements the anatomical and hemodynamic information provided by imaging techniques, making it an irreplaceable component of cardiac assessment.^22^ The ECG diagnostics of LVH currently is based primarily on the QRS voltage criteria. The current ECG diagnosis of LVH is largely based on QRS voltage criteria. This diagnostic approach is grounded in two main physiological principles. Firstly, LVH causes substantial alterations in QRS complex amplitudes, which correspond to the excitation of the hypertrophied ventricular walls, the specific lead showing the most prominent change depends on the location of the hypertrophy. Secondly, the increased myocardial tissue mass in severe LVH lowers its electrical impedance, thereby diminishing the overall impedance between the heart and the surface recording electrodes and contributing to the observed voltage increase. Several ECG criteria are available to assess LVH in adult hypertensive patients.^9^ However, affected by many factors such as intraventricular pressure, resistance of large blood vessels, abnormal cardiac position, anatomical relationship between the heart and the chest wall, and obesity, the diagnostic capability of ECG for LVH is compromised, resulting in a wide range of both sensitivities and specificities of ECG criteria for LVH.^20^ The number of ECG criteria for LVH implies that no single or explicit ECG pattern is predictive of an increase in LVM.

Our study successfully developed and validated a nomogram prediction model for LVH in children with primary hypertension. With BMI, ECG parameters R_1_+Sv_4_ and S_D_+Sv_4_ as independent predictors, this model significantly improves the diagnostic efficacy of ECG for LVH associated with primary hypertension in children. Studies indicate that increased fat mass is a key phenotypic characteristic of primary hypertension in adolescents, with overweight and obesity being its most typical clinical manifestations.^23^ In our study, the proportion of obese children reached 75.3%. Obesity-induced LVH is currently attributed to increased circulatory blood volume and sympathetic predominance, which lead to left ventricular dilation and wall thickening.^24^ Our previous study has also identified BMI as an independent risk factor for LVH in children with primary hypertension.^25^ However, in obese children, the increased thickness of the chest wall fat layer elevates the electrical impedance of thoracic tissues, which attenuates cardiac electrical signals transmitted to the body surface. Consequently, even in the presence of significant LVH, the QRS complex voltage may fail to meet conventional diagnostic criteria. Traditional ECG voltage standards do not correct for this confounding effect, resulting in low diagnostic sensitivity and false-negative diagnoses of LVH. The key innovation of our study lies in transforming this recognized confounder into a quantifiable corrective factor. By integrating BMI systematically into the ECG diagnostic model for LVH, we quantitatively account for voltage attenuation, thereby aligning the model more closely with the underlying pathophysiology. This approach refines traditional criteria, enabling the effective identification of obese patients with LVH who would otherwise be missed, significantly improves diagnostic sensitivity in the obese population, and enhances the the clinical utility of the ECG diagnostic model in hypertensive populations.

Based on a synthesis of previous research in both adults and children, our study incorporated a total of 22 ECG parameters for the diagnosing of LVH in children. The results showed that R_I_ + S_V4_ and S_D_ + S_V4_ were identified as independent predictors of LVH. Pediatric ECG parameters are prone to influence by individual variations such as age, height, weight, chest wall fat thickness, and cardiac axis (e.g., vertical or horizontal heart). Consequently, individual ECG indicators (e.g., R_Ⅰ_, R_aVL_, S_V_₄, S_Ⅲ_) generally exhibit limited sensitivity for diagnosing LVH, and criteria derived from adult populations have constrained applicability. Both R_I_ + S_V4_ and S_D_ + S_V4_ are composite voltage indices based on the standard 12-lead ECG. The R_I_ + S_V4_ index captures electrophysiological changes associated with anterolateral LVH, whereas S_D_ + S_V4_ may better align with pediatric cardiac anatomical positioning. By integrating these two indices, it is possible to capture electrophysiological abnormalities related to LVH from different perspectives, allowing them to mutually correct inherent biases and thereby enhance diagnostic sensitivity.

Compared with previously commonly used clinical models for LVH diagnosis^26,27^, Verified by NRI and IDI, the nomogram model constructed in this study demonstrates superior performance. Moreover, its AUC is significantly higher than that of traditional models, enabling more accurate identification of LVH cases among children with primary hypertension. In addition, the model is easy to operate. The required BMI is a routine physical examination indicator, R_1_+Sv_4_ and S_D_+Sv_4_ can be quickly obtained through standard 12-lead ECG without complex detection equipment. It is suitable for promotion and application in medical institutions at all levels, providing a reliable and convenient tool for the early screening of LVH related to primary hypertension in children, and holds great clinical significance for the early prevention and intervention of cardiovascular diseases in hypertensive children.

## Limitation

The study has several limitations that should be addressed in future research. Firstly, this was a single-center retrospective design, which may limit the model’s external validity, Multicenter prospective studies are needed to validate the model in more diverse cohorts. Secondly, the cohort included 79.68% males, which may reflect a higher prevalence of primary hypertension in boys, future studies should aim for more balanced gender representation to confirm whether the model performs consistently across sexes.Thirdly, the study focused on diagnostic performance rather than long-term outcomes. Evaluating whether the model can predict long-term cardiovascular risk in hypertensive children with LVH would further strengthen its clinical value.

## Conclusion

In conclusion, the nomogram model incorporating BMI, R_I_ + S_V4_, and S_D_ + S_V4_ significantly improves the ECG diagnosis of LVH in children with primary hypertension. This model demonstrates favorable discrimination, accuracy, and clinical applicability, outperforming existing alternatives and providing a reliable tool for the early detection of LVH in pediatric population.

## Data Availability

The datasets generated during and/or analyzed during the current study are available from the corresponding author on reasonable request.

## Abbreviations

LVH: Left ventricular hypertrophy
ECG: Electrocardiography
LVMI: Left ventricular mass index
RWT: Relative left ventricular wall thickness
ECHO: Echocardiography
LASSO: Least absolute shrinkage and selection operator
AUC: Area under the curve
ROC: Receiver operating characteristic
DCA: Decision curve analysis
NRI: Net reclassification improvement
IDI: Integrated discrimination improvement
BMI: Body mass index
TOD: Target organ damage
SBP: Systolic blood pressure
DBP: Diastolic blood pressure
LVIDd: Left ventricular internal dimension
IVST: Interventricular septal thickness
LVPWT: Left ventricular posterior wall thickness
LVM: Left ventricular mass

## Acknowledgments

We are grateful to the staff of the cardiology department of Capital Center for Children’s Health, Capital Medical University for support in data collection.

## Authors’ contributions

YL1 and LS conceived of the study. YL1 performed literature review. YL1 and YYL carried out data collection. YL1, LS, YXC, YL2, TZ and MB analyzed the data. YL1, YXC, and YL2 performed data interpretation. All authors (YL1, YXC, YL2, TZ, MB, YYL and LS) were involved in manuscript drafting and agreed with the submitted version of this manuscript.

## Declarations

**Ethics approval** This study had approval from the Ethic Committee of the Capital Center for Children’s Health, Capital Medical University (No: SHERLL2025011). Each child’s parent or legal guardian/next of kin provided signed informed consent. This study was performed in line with the principles of the Declaration of Helsinki and its later amendments.

**Consent to participate** All participants gave their written consent to participate.

**Consent for publication** Not applicable.

**Competing interests** The authors declare no competing interests.

